# A prospective study of asymptomatic SARS-CoV-2 infection among individuals involved in academic research under limited operations during the COVID-19 pandemic

**DOI:** 10.1101/2021.11.17.21266367

**Authors:** Audrey Pettifor, Bethany L. DiPrete, Bonnie E. Shook-Sa, Lakshmanane Premkumar, Kriste Kuczynski, Dirk Dittmer, Allison Aiello, Shannon Wallet, Robert Maile, Joyce Tan, Ramesh Jadi, Linda Pluta, Aravinda M. de Silva, David J. Weber, Min Kim, Arlene C. Seña, Corbin D. Jones

## Abstract

**Background:** Early in the pandemic, transmission risk from asymptomatic infection was unclear making it imperative to monitor infection in workplace settings. Further, data on SARS-CoV-2 seroprevalence within university populations has been limited.

**Methods:** We performed a longitudinal study of University research employees on campus July-December 2020. We conducted questionnaires on COVID-19 risk factors, RT-PCR testing, and SARS-CoV-2 serology using an in-house spike RBD assay, laboratory-based Spike NTD assay, and standard nucleocapsid platform assay. We estimated prevalence and cumulative incidence of seroconversion with 95% confidence intervals using the inverse of the Kaplan-Meier estimator.

**Results:** 910 individuals were included in this analysis. At baseline, 6.2% (95% CI 4.29-8.19) were seropositive using the spike RBD assay; four (0.4%) were seropositive using the nucleocapsid assay, and 44 (4.8%) using the Spike NTD assay. Cumulative incidence was 3.61% (95% CI: 2.04-5.16). Six asymptomatic individuals had positive RT-PCR results.

**Conclusions:** Prevalence and incidence of SARS-CoV-2 infections was low; however differences in target antigens of serological tests provided different estimates. Future research on appropriate methods of serological testing in unvaccinated and vaccinated populations is needed. Frequent RT-PCR testing of asymptomatic individuals is required to detect acute infections, and repeated serosurveys are beneficial for monitoring subclinical infection.

## INTRODUCTION

The SARS-CoV-2 pandemic caused significant disruptions to research programs across university campuses. In March of 2020, most researchers, support staff, and trainees were sent home as non-essential research activities were halted. Super-spreader events early in the pandemic had engendered concern about the safety of working in-person, due to the possibility of asymptomatic infections among individuals and the potential for spread of COVID-19 within the workplace [1, 2]. Thus, only essential research staff remained on campus throughout the early months of the pandemic. However, beginning mid-2020, many universities started bringing research staff back to campuses following new safety protocols, with different approaches to testing for SARS-CoV-2 to identify infections in the workplace.

Approximately 30% of unvaccinated individuals are estimated to have asymptomatic infection with SARS-CoV-2 [3], and asymptomatic individuals may account for approximately 25% of transmissions [4]. Individuals with asymptomatic infection appear to harbor similar viral load levels to those of symptomatic individuals. However, asymptomatic individuals are likely to shed for fewer days than symptomatic individuals [5]. Shorter duration of viral shedding may have implications for the effectiveness of regular SARS-CoV-2 testing strategies to detect acute infections among asymptomatic individuals in the workplace. Furthermore, pre-symptomatic individuals are known to be infectious, with viral shedding demonstrated 48 hours prior to symptom onset [6].

The primary objective of this study was to describe the extent of SARS-CoV-2 seroprevalence and incidence among the research community at an academic university from July to December 2020.

## METHODS

### Eligibility and Recruitment

We conducted a longitudinal cohort study of research faculty, staff, and students who were coming to University of North Carolina at Chapel Hill (UNC-CH) campus at least one day per week between July and December 2020. Employees or students who conducted or supported research activities, reported coming to campus at least one day a week, and were age 18 or older were eligible to enroll. All individuals who received funding from research grants were invited to participate.

Screening and enrollment took place online via a REDCap survey. Consent was provided electronically. After enrollment, the study team scheduled the participant for an in-person appointment. The day before the appointment, the participant received an invitation to complete an online questionnaire and assessed for symptoms of SARS-CoV-2. Anyone with symptoms was instructed to call their healthcare provider, follow UNC-CH guidelines for COVID-19 testing, and to re-schedule their study visit. On the day of the visit, screening procedures for COVID-19 were repeated.

Participants who also enrolled in a COVID-19 vaccine trial during the study period were excluded from the analyses.

### Questionnaire

Participants completed the online survey at baseline and months 1 and 3 after enrollment. The survey collected information on symptoms, compliance with COVID-19 public health measures at work and in the community, perceived safety at work, and mental health. Participants were screened for symptoms of depression using the Patient Health Questionnaire-2 (PHQ-2) [7] and for symptoms of anxiety using the Generalized Anxiety Disorder-2 (GAD-2) [8].

### Sample Collection

At each in-person study visit at baseline and months 1 and 3 after enrollment, participants underwent RT-PCR testing for SARS-CoV-2 using a self-collected, observed mid-turbinate nasal swab, and serology testing for SARS-CoV-2 using blood draws (5 mL). At the first visit, participants were guided on how to properly collect the nasal swab. Participants watched an instructional video on self-collection and were provided an instruction sheet.

Participants were invited to follow-up in-person visits approximately 1 and 3 months after the initial baseline visit. Similar procedures were conducted at month 1 and 3 with one exception: at month 1, participants were provided a Tasso device (Tasso, Inc., Seattle, WA, USA) for self-administered blood collection (30-80 μl), which they could take home and return at a later time to test for SARS-CoV-2 antibodies. In between monthly visits, all participants were asked to conduct self-collected nasal swabs approximately every 2 weeks for RT-PCR testing for SARS-CoV-2. To reduce in-person contact time, participants were provided the option to pick up nasal swab kits and drop them off after specimen collection for all visits after baseline.

### Laboratory Assays

#### Sample Accessioning and Preprocessing

All samples, blood or nasal, were accessioned and pre-processed within 24 hours of collection at the DELTA Translational Core. Multiple aliquots were generated from each sample, material permitting. Individual aliquots were then directed to specific assays and any residual material was banked at the UNC-CH Biospecimen Processing Center.

#### Real-Time Reverse Transcription Polymerase Chain Reaction (RT-PCR) Assay

Mid-turbinate nasal swabs collected from individuals participating in the study were analyzed using RT-PCR for the detection of nucleic acid from SARS-CoV-2. The RT-PCR assay used for this study was based on the initial assay implemented for the UNC Respiratory Diagnostic Clinic by the Clinical Microbiology and Molecular Microbiology Laboratories at UNC Hospitals. In brief, RNA was extracted using the Roche Diagnostics MagNA Pure MPC large volume isolation system (Roche Diagnostics, Indianapolis, IN, USA) and NucleoSpin Isolation kit (MACHEREY-NAGEL GmbH & Co. KG, Germany). Samples were then quantified, reverse transcribed (SuperScript 3, ThermoFisher, USA) on a thermocycler. Using primers based on the Respiratory Diagnostic Clinic assay and human RNA control primers, amplicons were amplified and then quantified using a ThermoFisher QuantStudio 7 system. While based on the clinical assay, this RT-PCR assay did not have Emergency Use Authorization (EUA) from the U.S. Food and Drug Administration (FDA); therefore, all positive results were referred for confirmatory testing using an EUA-approved PCR test in a Clinical Laboratory Improvement Amendments (CLIA) approved laboratory 3-10 days after the research sample was collected.

#### Serological assays to detect SARS-CoV-2 antibodies

Serum IgG antibodies were analyzed using three serological assays. The first was the commercially available Abbott SARS-CoV-2 assay (Abbott, Chicago, IL, USA) to detect IgG antibodies to nucleocapsid antigen using a chemiluminescent microparticle immunoassay (CMIA), which had received an EUA. The reported sensitivity and specificity among those with confirmed SARS-CoV-2 is 100% and 99.6%, respectively; however, the test is not suitable for samples collected less than seven days after onset due to low sensitivity in the first week after onset. The CMIA provides qualitative detection of SARS-CoV-2 IgG antibodies on the Abbott Architect instrument. Results are reported as nonreactive or reactive. The Abbott IgG assay was used on samples collected at the baseline and month 3 visit.

In addition, two in-house Enzyme-Linked Immunosorbent Assays (ELISA) based on the receptor-binding domain (RBD) and the N-terminal domain (NTD) of the SARS-CoV-2 spike protein were performed as previously described [9, 10]. The sensitivity and specificity of the test were 98% and 100%, respectively, 9 days after symptom onset in patients with confirmed SARS-CoV-2 infection. Briefly, heat-inactivated serum samples collected at baseline, month 1, or month 3 were diluted at 1:40 in a TBS-based diluent buffer with 3% Bovine Serum Albumin (BSA), 0.05% Tween 20, and biotinylated spike RBD or NTD antigen at 1 μg/mL. After incubation for 1 hour at 37°C, antibodies bound to biotinylated antigen were captured onto a streptavidin-coated assay plate. The assay plate was washed, then a cocktail of horseradish peroxidase-conjugated secondary Goat Anti-Human IgG, IgA, and IgM secondary antibodies was used to measure antigen-specific total Ig. The optical density (OD) thresholds for seropositivity in the RBD (≥0.37) and NTD (≥0.27) assays were used based on reference panel performance.

### Statistical Analyses

To mitigate potential selection bias in the sample, study participants were weighted to the target population of approximately 5,019 UNC-CH researchers working on campus by gender, school of appointment (categorized as College of Arts & Sciences, School of Pharmacy, School of Global Public Health, School of Medicine, and other schools), role on campus (principal investigator (PI)/faculty, postdoctoral researcher, student, or research support personnel), race/ethnicity (Hispanic, non-Hispanic Asian, non-Hispanic Black, non-Hispanic white, non-Hispanic other race or 2 or more races), and age category (18-24, 25-34, 35-44, 45-54, 55-64, and 65+). Linear weight calibration was performed using the calibrate function in the R survey package [11]. Control totals for calibration came from the weighted results of a survey conducted among researchers in the summer of 2020, which asked researchers whether they were working on campus or if their job responsibilities required them to work on campus. The resulting weights ranged from 1.0 to 17.3, with a median weight of 5.2.

We calculated both unweighted and weighted seroprevalence at each study visit as the proportion of samples with a positive result, with 95% confidence intervals (CI). To analyze agreement between the spike RBD assay and (1) nucleocapsid assay or (2) spike NTD assay, we first compared prevalence estimates using each assay. We then examined concordance using McNemar’s test (*χ*^2^)and Cohen’s Kappa (κ). Using the spike RBD assay, we analyzed longitudinal binding results among participants who were (1) seropositive at baseline, and (2) seronegative at baseline who seroconverted during follow-up.

We estimated unweighted and weighted cumulative incidence of seroconversion among participants who were seronegative at baseline and did not enroll in a vaccine trial using the inverse of the Kaplan-Meier estimator. The date of seroconversion was defined as the mid-point between the last negative test and first positive test result, and participants were considered to be censored on the known date of withdrawal from the study or 30 days after their last sample.

Finally, we examined predictors of seroconversion using unweighted and weighted Cox proportional hazards models to estimate unadjusted hazard ratios (HR) and 95% confidence intervals (CI), using Schoenfeld’s residuals to test the proportional hazards assumption. Predictors considered were demographic and household characteristics, role on campus, symptoms of anxiety or depression, compliance with public health guidelines for COVID-19 in the workplace and in public, any reported symptoms in the last two weeks, and any reported travel within the past two weeks. For time-varying measures, we determined whether a participant had ever endorsed the measure prior to the first positive test for participants who seroconverted or end of follow-up for participants who remained negative. For all other measures, the baseline value was used.

All analyses were conducted in R version 4.0.2 (Vienna, Austria) [12].

### Ethical Approval

This study was approved by the Institutional Review Board at the University of North Carolina at Chapel Hill.

## RESULTS

There were 927 individuals enrolled in the study. Seven participants enrolled in a vaccine trial during follow-up and were excluded from analyses. Of the remaining 920 individuals, 910 (99%) had a baseline serology assay completed and were included in the analytic cohort. Table 1 displays unweighted and weighted characteristics of the participants included in analyses. The majority of participants were female (60%), non-Hispanic white (78%), had earned a graduate degree (61%), did not live with children in the house (69%), and did not live with an essential worker (68%). Half of participants were aged 18-35; one-third worked in a research support role on campus. The vast majority of participants reported being compliant with regard to mask use (workplace: 94%, public: 96%), physical distancing (workplace: 87%, public: 92%), and gathering in groups <10 people (workplace: 92%, public: 93%), and 87% reported that all of their coworkers were compliant about mask use in the office. Of the 910 participants who submitted a baseline sample, 825 (91%) provided at least one follow-up sample for serologic testing, and 677 (74%) had serologic testing completed at all three study visits. Seventeen percent withdrew prior to the final visit (Table 2).

**Table 1.** Baseline characteristics of the cohort of University research employees with a baseline serology result (N=910), unweighted and weighted to the target population of all University researchers.

|  | Overall - Unweighted<br>N (%) | Overall - Weighted<br>N (%; 95% CI) |
| --- | --- | --- |
| Age (years) |  |  |
| 18-25 | 170 (18.8) | 901 (18.4, 15.4-21.4) |
| 26-35 | 285 (31.5) | 1929 (39.4, 35.5-43.2) |
| 36-45 | 198 (21.9) | 916 (18.7, 15.8-21.6) |
| 46-55 | 127 (14.0) | 553 (11.3, 9.0-13.6) |
| >55 | 124 (13.7) | 597 (12.2, 9.8-14.6) |
| Gender |  |  |
| Men | 356 (39.2) | 2364 (48.0, 44.1-51.8) |
| Women | 544 (59.8) | 2518 (51.1, 47.2-55.0) |
| Other | 9 (1.0) | 45 (0.9, 0.1- 1.7) |
| Race/Ethnicity |  |  |
| Hispanic | 54 (5.9) | 292 (5.9, 4.2- 7.7) |
| Asian | 95 (10.4) | 1036 (21.0, 17.3-24.8) |
| Black | 29 (3.2) | 268 (5.4, 3.3- 7.5) |
| White | 712 (78.2) | 3158 (64.1, 60.0-68.1) |
| Other/Multiple races | 20 (2.2) | 176 (3.6, 1.9- 5.2) |
| Education Level |  |  |
| Some college or less | 45 (5.0) | 181 (3.7, 2.3- 5.1) |
| College | 312 (34.3) | 1634 (33.2, 29.5-36.8) |
| Graduate | 552 (60.7) | 3112 (63.2, 59.5-66.9) |
| Role on Campus |  |  |
| Healthcare | 142 (15.7) | 657 (13.4, 10.9-15.9) |
| PI<sup>a</sup> + Teaching Faculty | 166 (18.3) | 925 (18.8, 15.9-21.8) |
| Postdocs | 98 (10.8) | 679 (13.8, 11.0-16.7) |
| Research Support | 283 (31.3) | 1170 (23.8, 20.6-27.0) |
| Students | 216 (23.9) | 1476 (30.1, 26.4-33.7) |
| Living with Essential Worker |  |  |
| Yes - First responder | 111 (12.3) | 559 (11.4, 9.1-13.8) |
| Yes - Not first responder | 177 (19.6) | 826 (16.9, 14.1-19.7) |
| Other | 614 (68.1) | 3507 (71.7, 68.3-75.1) |
| Children in Household |  |  |
| Living with children | 283 (31.1) | 1517 (30.8, 27.2-34.3) |
| Mental Health & Risk Perception |  |  |
| Has depressive symptoms | 115 (12.6) | 687 (13.9, 11.2-16.7) |
| Has anxiety symptoms | 175 (19.2) | 906 (18.4, 15.4-21.3) |
| Does not feel safe in workplace | 75 (8.3) | 390 (7.9, 5.7-10.1) |
| Compliance Measures |  |  |
| Gathered with people outside of home | 283 (31.1) | 1426 (28.9, 25.5-32.4) |
| Spent time with others INSIDE, no mask | 364 (40.0) | 1911 (38.8, 35.0-42.5) |
| Workplace: Solo office | 255 (28.0) | 1268 (25.7, 22.5-29.0) |
| Workplace: Compliant mask use | 849 (94.1) | 4623 (94.4, 92.7-96.1) |
| Workplace: Maintained physical distancing | 780 (86.5) | 4221 (86.1, 83.3-88.8) |

| **Table 1 cont.** | Overall - Unweighted<br>N (%) | Overall - Weighted<br>N (%; 95% CI) |
| --- | --- | --- |
| Workplace: Only met in groups <10 | 827 (91.6) | 4469 (91.1, 88.9-93.4) |
| Workplace: 100% of coworkers wear masks | 785 (87.1) | 4275 (87.6, 85.1-90.1) |
| Workplace: Never ate indoors | 704 (77.9) | 3826 (78.0, 74.8-81.2) |
| Public: Compliant mask use | 874 (96.4) | 4759 (96.7, 95.5-98.0) |
| Public: Maintained social distancing | 828 (91.5) | 4540 (92.2, 90.3-94.2) |
| Public: Only met in groups <10 | 839 (92.7) | 4526 (92.1, 90.1-94.2) |
| Travel |  |  |
| Any travel | 462 (50.8) | 2450 (49.7, 45.8-53.5) |
| Travel out of state | 99 (10.9) | 544 (11.0, 8.7-13.4) |
| Travel using public transportation | 29 (3.2) | 161 (3.3, 1.9- 4.6) |
| Any COVID-19 Symptoms |  |  |
| Yes, $\geq 1$ symptom in last 2 weeks | 170 (18.7) | 924 (18.8, 15.7-21.8) |
| Missing data: age (n=6), gender (n=1), education (n=1), role on campus (n=5), living with essential worker (n=8), feeling safe in the workplace (n=3), workplace: mask use (n=8), workplace: physical distancing (n=8), workplace: met in groups <10 (n=7), public: mask use (n=3), public: physical distancing (n=5), public: met in groups <10 (n=5) |  |  |
| <sup>a</sup> PI, principal investigator |  |  |

**Table 2.** Cumulative seroprevalence over the study period using the anti-spike RBD assay.

|  | N | Unweighted<br>(95% CI) | Weighted<br>(95% CI) |
| --- | --- | --- | --- |
| Baseline | 910 | 5.38 (3.92-6.85) | 6.24 (4.29-8.19) |
| Month 1 | 751 | 8.66 (6.64-10.67) | 9.21 (6.74-11.68) |
| Month 3 | 751 | 8.92 (6.88-10.96) | 10.07 (7.41-12.72) |

Cumulative seroprevalence over the study period based on the unweighted study population (N=910) and weighted to the target population of all University researchers

There were very few active SARS-CoV-2 infections detected using the research RT-PCR among asymptomatic participants; five participants tested positive at baseline and one tested positive at the first bi-weekly mid-turbinate nasal swab test. An additional sample was collected in the clinical lab between 3 and 10 days after the research sample was collected, and none of these participants had a positive confirmatory RT-PCR test result. One participant who was tested positive by the research PCR had a positive serological result. The median time between nasal swabs during the study was 22 days (Q1-Q3: 14-40 days).

At baseline, the prevalence of SARS-CoV-2 using the in-house spike RBD ELISA IgG assay was 6.24% (95% CI 4.29-8.19) weighted to the target population of all researchers and 5.38% (95% CI 3.92-6.85) unweighted (n=49, Table 2). Fig 1A displays the distribution of OD values among study participants at baseline. In comparison, four participants (0.4%) were seropositive at baseline using the nucleocapsid assay (Comparing to RBD: McNemar’s test *χ*^2^=40.20, p<0.001; Cohen’s Kappa κ=0.10), and 44 (4.8%) were seropositive using the Spike NTD ELISA assay (Comparing to RBD: *χ*^2^=0.59, p=0.44; κ=0.69) (Table 3). There was substantial concordance between Spike RBD and Spike NTD serological assays (Fig 1B). There was poor concordance between the Spike RBD and Abbott nucleocapsid serological assays (Fig 1C). Cumulative prevalence of SARS-CoV-2 throughout the study using the Spike RBD ELISA was 8.92% (95% CI 6.88-10.96) unweighted and 10.07% (95%CI 7.41-12.72) weighted (Table 2).

**Table 3.** Comparisons of baseline serology results based on the nucleocapsid IgG (Abbott) and spike NTD total Ig assays versus the spike RBD total Ig assay.

| | Positive | Negative<sup>a</sup> | McNemar's Test<br>$\chi^2$ (p-value) | Cohen's Kappa<br>$\kappa$ |
| --- | --- | --- | --- | --- |
| Spike RBD Assay | 49 (5.4%) | 861 (94.6%) | - | - |
| Abbott Assay<sup>b</sup> | 4 (0.4%) | 901 (99.6%) | 40.20 (p<0.001) | 0.10 |
| Spike NTD Assay | 44 (4.8%) | 866 (95.2%) | 0.59 (p=0.44) | 0.69 |

<sup>a</sup> Inconclusive results treated as negative for the spike RBD assay
<sup>b</sup> Abbott SARS-CoV-2 results missing for 5 participants at baseline

**Fig 1.**
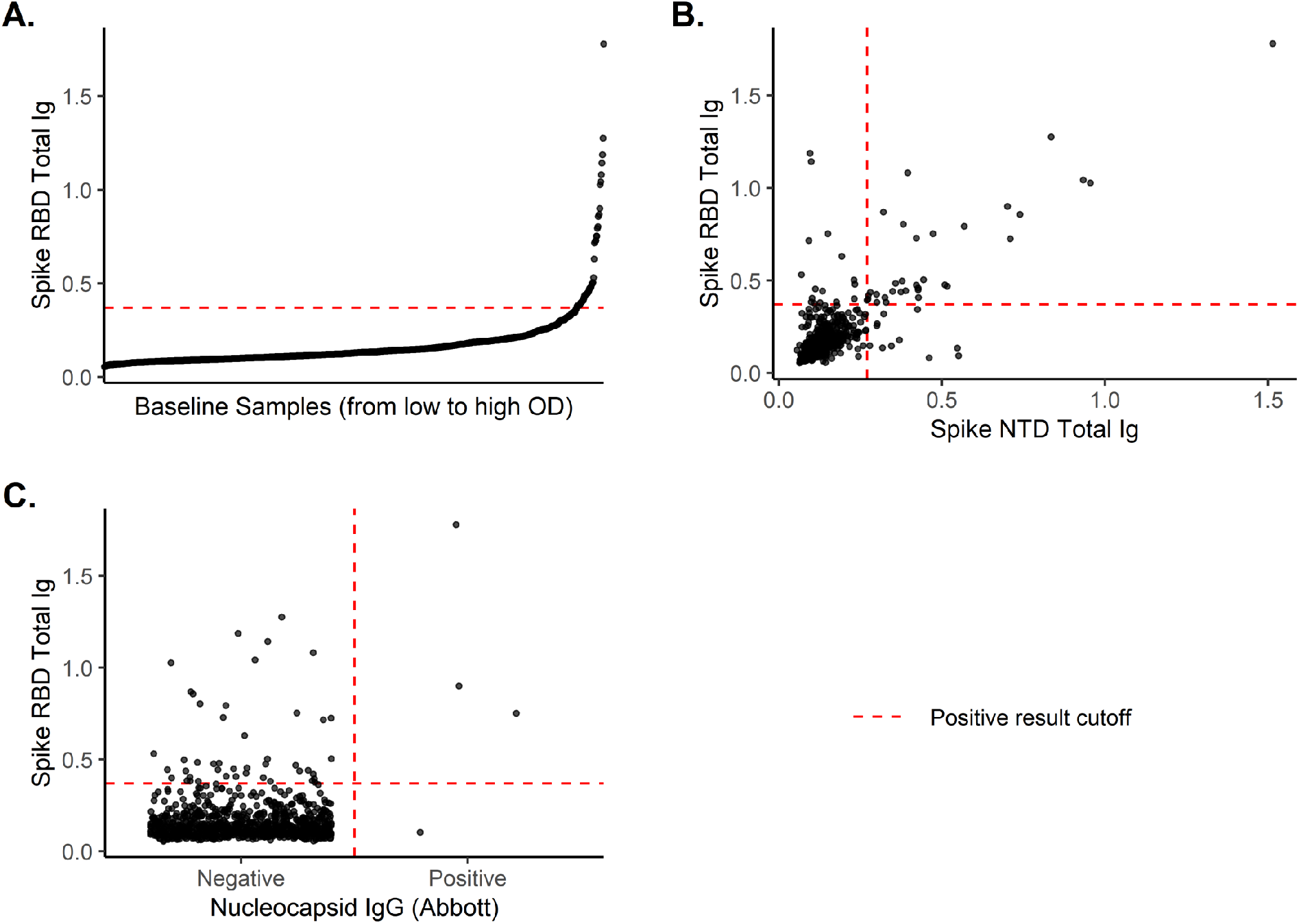
Baseline serological results showing (A) the reactivity to spike RBD and its concordance with (B) Spike NTD and (C) Nucleocapsid IgG (Abbott) assays.

Figure 2 displays longitudinal binding results among participants who were (A) seronegative at baseline and seroconverted by their month 1 sample (N=22), and (B) seronegative at baseline and seroconverted by their month 3 sample (N=8). Cumulative incidence of seroconversion among the 861 participants who were seronegative at baseline and did not enroll in a vaccine trial was 3.64% (95% CI: 2.35-4.91) unweighted and 3.61% (95% CI: 2.04-5.16) weighted.

**Fig 2.**
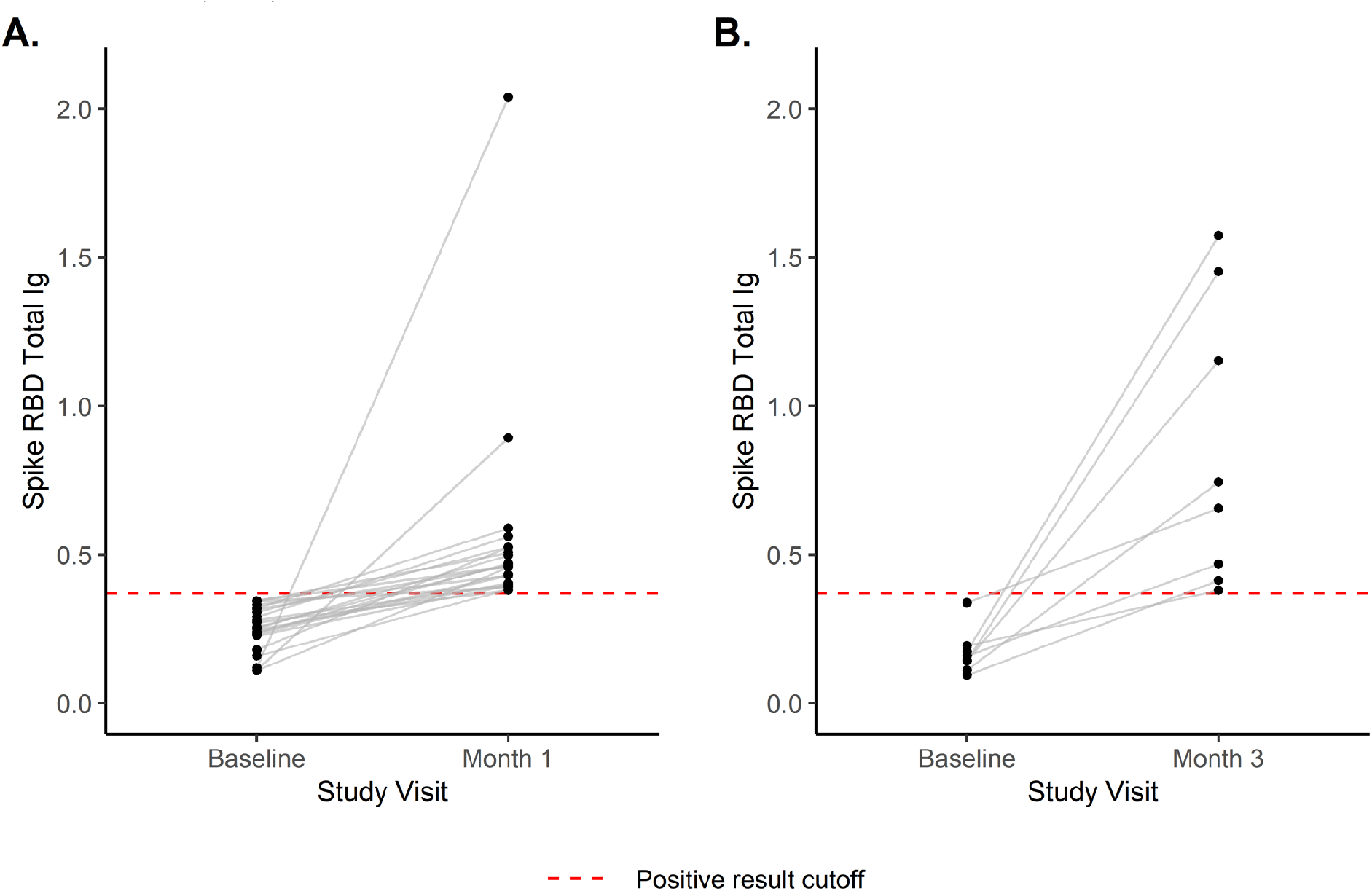
Longitudinal spike RBD binding results among individuals who were seronegative at baseline and (A) seroconverted by the month 1 visit (N=22), and (B) seroconverted by the month 3 visit (N=8)

Table 4 displays unadjusted predictors of seroconversion. The strongest predictors of seroconversion were meeting in groups of 10 or more at work (Unweighted HR 2.97, 95% CI: 1.21-7.27) or in public (Unweighted HR 2.74, 95% CI: 1.05-7.16). Individuals in PI/teaching faculty, post-doctoral, research support, and student roles were all less likely than research personnel with face-to-face patient contact to seroconvert during follow-up, although none of these associations were significant. There was no clear observed relationship between participants reporting any symptoms at any point prior to their positive test and seroconversion.

**Table 4.** Bivariate associations between predictors and seroconversion among participants who were seronegative at baseline, unweighted and weighted to the target population of all University

|  | Seroconverted<br>N (%) | Unweighted HR<sup>a</sup><br>(95% CI) | Weighted HR<br>(95% CI) |
| --- | --- | --- | --- |
| Age (years) |  |  |  |
| 18-25 | 5 (3.2) | - | - |
| 26-35 | 9 (3.4) | 1.03 (0.35-3.09) | 0.82 (0.25-2.70) |
| 36-45 | 7 (3.7) | 1.13 (0.36-3.55) | 0.67 (0.19-2.40) |
| 46-55 | 7 (5.7) | 1.76 (0.56-5.54) | 0.96 (0.27-3.36) |
| >55 | 2 (1.7) | 0.52 (0.10-2.66) | 0.27 (0.04-1.89) |
| Gender |  |  |  |
| Men | 14 (4.2) | - | - |
| Women | 15 (2.9) | 0.68 (0.33-1.42) | 1.14 (0.47-2.77) |
| Race/Ethnicity |  |  |  |
| Non-white or Hispanic | 7 (3.8) | - | - |
| White, non-Hispanic | 23 (3.4) | 0.86 (0.37-2.01) | 0.72 (0.29-1.84) |
| Education Level |  |  |  |
| Some college or less | 2 (5.1) | - | - |
| College | 9 (3.0) | 0.57 (0.12-2.62) | 0.33 (0.05-2.21) |
| Graduate | 19 (3.6) | 0.68 (0.16-2.91) | 0.38 (0.06-2.40) |
| Role on Campus |  |  |  |
| Healthcare | 9 (6.7) | - | - |
| PI<sup>b</sup> + teaching faculty | 3 (1.9) | 0.28 (0.08-1.04) | 0.18 (0.04-0.84) |
| Postdocs | 3 (3.2) | 0.47 (0.13-1.75) | 0.69 (0.17-2.82) |
| Research support | 10 (3.8) | 0.56 (0.23-1.39) | 0.45 (0.14-1.42) |
| Students | 5 (2.5) | 0.37 (0.13-1.11) | 0.34 (0.10-1.14) |
| Living with Essential Worker |  |  |  |
| Other | 20 (3.5) | - | - |
| Living with essential, first responder | 4 (3.8) | 1.09 (0.37-3.18) | 0.86 (0.25-2.95) |
| Living with essential, not first responder | 6 (3.5) | 1.02 (0.41-2.54) | 1.20 (0.42-3.37) |
| Children in Household |  |  |  |
| Not living with children | 22 (3.7) | - | - |
| Living with children | 8 (3.0) | 0.81 (0.36-1.81) | 0.54 (0.20-1.43) |
| Depressive Symptoms |  |  |  |
| Does not have depressive symptoms | 25 (3.3) | - | - |
| Has depressive symptoms | 5 (4.7) | 1.44 (0.55-3.76) | 1.52 (0.49-4.68) |
| Anxiety |  |  |  |
| Does not have anxiety symptoms | 23 (3.3) | - | - |
| Has anxiety symptoms | 7 (4.3) | 1.31 (0.56-3.05) | 1.42 (0.55-3.65) |
| Feels safe at work |  |  |  |
| Feels safe in workplace | 28 (3.6) | - | - |
| Does not feel safe in workplace | 2 (2.9) | 0.84 (0.20-3.52) | 0.68 (0.13-3.51) |
| **Table 4 cont.** | Seroconverted<br>N (%) | Unweighted HR<sup>a</sup><br>(95% CI) | Weighted HR<br>(95% CI) |
| Primary workplace: Solo office |  |  |  |
| No | 24 (3.9) | - | - |
| Yes | 6 (2.5) | 0.63 (0.26-1.54) | 0.47 (0.17-1.35) |
| Workplace: Worn a Mask |  |  |  |
| Compliant | 28 (3.5) | - | - |
| Not compliant | 2 (3.8) | 1.08 (0.26-4.55) | 0.20 (0.05-0.88) |
| Workplace: Only met in groups <10 |  |  |  |
| Compliant | 24 (3.1) | - | - |
| Not compliant | 6 (8.7) | 2.97 (1.21-7.27) | 3.10 (1.03-9.36) |
| Workplace: Percentage of coworkers wearing masks |  |  |  |
| 100% | 21 (3.5) | - | - |
| <100% | 9 (3.6) | 1.01 (0.46-2.20) | 0.80 (0.31-2.07) |
| Public: Worn a Mask |  |  |  |
| Compliant | 27 (3.3) | - | - |
| Not compliant | 3 (9.1) | 2.81 (0.85-9.26) | 1.37 (0.29-6.53) |
| Public: Maintained Social Distancing |  |  |  |
| Compliant | 26 (3.3) | - | - |
| Not compliant | 4 (5.6) | 1.68 (0.59-4.82) | 0.98 (0.27-3.64) |
| Public: Only met in groups <10 |  |  |  |
| Compliant | 25 (3.1) | - | - |
| Not compliant | 5 (8.2) | 2.74 (1.05-7.16) | 3.63 (1.22-10.80) |
| Gone out & gathered with people not in your household |  |  |  |
| No | 24 (3.6) | - | - |
| Yes | 6 (3.0) | 0.83 (0.34-2.04) | 0.43 (0.15-1.29) |
| Spent time with friends, neighbors, relatives INSIDE, no mask |  |  |  |
| No | 16 (2.9) | - | - |
| Yes | 14 (4.4) | 1.52 (0.74-3.12) | 1.48 (0.62-3.52) |
| Any COVID-19 Symptoms |  |  |  |
| No | 18 (3.7) | - | - |
| Yes | 12 (3.2) | 0.81 (0.39-1.67) | 0.92 (0.38-2.18) |
| Any travel |  |  |  |
| No | 7 (3.5) | - | - |
| Yes | 23 (3.5) | 0.94 (0.40-2.20) | 1.98 (0.67-5.85) |

<sup>a</sup> HR=Hazard Ratio
<sup>b</sup> PI = Principal Investigator

## DISCUSSION

In this cohort of University research staff and students who provided routine asymptomatic SARS-COV-2 testing from June-December 2020, RT-PCR detected few positive results (n=6), while serology identified an incidence of 3.6% (n= 30) over the approximate 3 month study duration. Baseline seroprevalence ranged from 0.5% using the nucleocapsid Abbott SARS-CoV-2 IgG assay to 5.4% using an in-house spike RBD IgG ELISA assay. While this university cohort engaged in research generally reported good compliance with COVID-19 public health guidelines, gathering in groups of 10 or more at work and in public was associated with seroconversion.

Viral shedding in asymptomatic individuals varies greatly but has been shown to last for fewer days compared to those with symptomatic infections [13]. In one study with daily nasal swab testing, viral load was detectable on average for 6.7 days in asymptomatic individuals but the 95% CI ranged from 3.9-9.2 days [14]. This may have accounted for the fact that none of our positive research PCR tests were positive on the confirmatory test, which on average was collected 3-10 days after the research PCR and potentially later in the viral shedding trajectory. The short duration of viral shedding in asymptomatic individuals and our finding that approximately bi-weekly testing with RT-PCR (collected on average every 22 days) likely missed infections that occurred during follow-up suggests that a more frequent testing schedule is needed to detect asymptomatic infections. Furthermore, the sensitivity of RT-PCR testing to detect asymptomatic infections is not clear, and lower viral load in asymptomatic individuals may have resulted in false negative RT-PCR results even despite testing within the window of viral shedding.

We found very low concordance (almost a 5x difference in seroprevalence) between serological testing modalities comparing anti-nucleocapsid with the Abbott platform and our lab-based ELISA for RBD. Our data strongly suggest that the target antigen of the selected assay is likely critical for detection of mild and asymptomatic cases and may account for the differences that we observed. Evidence suggests that assays targeting the spike protein outperform assays targeting the nucleocapsid in individuals with low levels of antibodies to SARS-CoV-2 [15, 16]. Specifically, a longitudinal study of healthcare workers in the United Kingdom found that anti-nucleocapsid antibodies wane within months of infection, with more rapid declines in asymptomatic individuals, whereas anti-spike IgG levels were sustained for up to six months [17]. Taken together, this evidence supports our finding of poor concordance between the spike RDB IgG and the assay targeting the nucleocapsid, with far fewer asymptomatic infections detected using the anti-nucleocapsid assay than the in-house anti-spike RBD assay. After vaccination, however, assays based on spike antigen are unsuitable for detecting SARS-CoV-2 infection [9]. Hence, there is an urgent need for developing more sensitive and robust methods based on a non-spike antigen to detect individuals who experienced asymptomatic or mild infection to control community spread.

Seroprevalence in our study population of university employees engaged in research on campus was likely lower than the general population of North Carolina. A study of two cohorts of asymptomatic individuals presenting to healthcare clinics in North Carolina from March through June 2020 found 0.7% and 0.8% of asymptomatic participants were positive in each cohort using the Abbott nucleocapsid assay, with evidence of a rising trend in seropositivity over time [18]. A nationwide analysis estimated seroprevalence in North Carolina ranged from 2.5% end of July 2020 to 6.8% by late September 2020 using the Abbott SARS-CoV-2 nucleocapsid assay [19]. However, the incidence of SARS-CoV-2 infection during follow-up in our study remained low despite rising case numbers in the state Carolina during the same months [20].

In this cohort, we relied on participants’ self-report of lack of symptoms associated with SARS-CoV-2 infection. Additionally, the study questionnaire asked participants to self-report behaviors related to compliance with COVID-19 public health guidelines, symptoms of COVID-19 within the past two weeks, travel, and other risk factors. These measures may have been subject to recall and social desirability bias, particularly questions related to COVID-19 compliance. The estimates of seroprevalence reported in this study were not adjusted for sensitivity and specificity of the assays because sensitivity and specificity estimates of the assays were based on symptomatic populations and estimates of assay performance among asymptomatic individuals was unavailable. Seventeen percent of study participants withdrew prior to study completion; therefore, our estimates of seroconversion may be subject to selection bias. Finally, while we weighted our analyses to the target population of all research staff, faculty, and students on campus based on available covariates and found comparable results, this population of university employees may not be fully representative of the target population due to unmeasured differences between the cohort and the target population.

## Conclusions

In conclusion, the prevalence and incidence of SARS-CoV-2 infection among employees and students involved in academic research under restricted operations in 2020 was relatively low. There was good compliance with COVID-19 prevention measures in the workplace in our study. We observed that different serological tests can provide very different estimates of seroprevalence. This may be due to sensistivity differences, asymptomatic cases, or viral target for immunogenicity. Future research on the most appropriate methods of serological testing in both unvaccinated and vaccinated populations is needed. Our findings suggest that frequent RT-PCR testing of asymptomatic individuals is required to detect acute infections, and repeated serosurveys are beneficial for detecting SARS-CoV-2 infection in predominantly asymptomatic populations.

## Data Availability

Deidentified data is available on request for any interested researchers to allow replication of results provided all ethical requirements are met.

## ACKNOWLEDGMENTS

Melissa Miller, Amir Barzin, Mike Cohen, Joe Eron, Andy Johns, Ryan McNamara, Femi Cleola S Villamor, Chuck Scheier, Greg Bowan, Asher Thomas, Austin Johnson, Piotr Mieczkowski ,Tara Skelly, Brooke Bullington, Michael Jetsupphasuk, Cailtlin Cassidy DELTA Translational Core: William Lovell, Olivia Mitchem, Dominique Burgess, Jessica Suggs General Oral Health Clinical Research Unit (GoHealth): Carol Culver, Wendy Lamm, ST Phillips

## REFERENCES

1. Lewis D. Superspreading drives the COVID pandemic — and could help to tame it. Nature. 2021;590(7847):544–6. doi: doi:10.1038/d41586-021-00460-x.

2. Lemieux JE, Siddle KJ, Shaw BM, Loreth C, Schaffner SF, Gladden-Young A, et al. Phylogenetic analysis of SARS-CoV-2 in Boston highlights the impact of superspreading events. 2021. doi: 10.1126/science.abe3261.

3. Oran D, Topol E. Prevalence of Asymptomatic SARS-CoV-2 Infection : A Narrative Review. Annals of internal medicine. 2020;173(5). doi: 10.7326/M20-3012. PubMed PMID: 32491919.

4. Johansson M, Quandelacy T, Kada S, Prasad P, Steele M, Brooks J, et al. SARS-CoV-2 Transmission From People Without COVID-19 Symptoms. JAMA network open. 2021;4(1). doi: 10.1001/jamanetworkopen.2020.35057. PubMed PMID: 33410879.

5. Cevik M, Tate M, Lloyd O, Maraolo A, Schafers J, Ho A. SARS-CoV-2, SARS-CoV, and MERS-CoV viral load dynamics, duration of viral shedding, and infectiousness: a systematic review and meta-analysis. The Lancet Microbe. 2021;2(1). doi: 10.1016/S2666-5247(20)30172-5. PubMed PMID: 33521734.

6. Kim S, Jeong H, Yu Y, Shin S, Kim S, Oh T, et al. Viral kinetics of SARS-CoV-2 in asymptomatic carriers and presymptomatic patients. International journal of infectious diseases : IJID : official publication of the International Society for Infectious Diseases. 2020;95. doi: 10.1016/j.ijid.2020.04.083. PubMed PMID: 32376309.

7. Kroenke K, Spitzer RL, Williams JB. The Patient Health Questionnaire-2: validity of a two-item depression screener. Medical care. 2003;41(11):1284-92. Epub 2003/10/30. doi: 10.1097/01.MLR.0000093487.78664.3C. PubMed PMID: 14583691.

8. Spitzer RL, Kroenke K, Williams JB, Lowe B. A brief measure for assessing generalized anxiety disorder: the GAD-7. Archives of internal medicine. 2006;166(10):1092-7. Epub 2006/05/24. doi: 10.1001/archinte.166.10.1092. PubMed PMID: 16717171.

9. Narowski TM, Raphel K, Adams LE, Huang J, Vielot NA, Jadi R, et al. SARS-CoV-2 mRNA Vaccine Induces Robust Specific and Cross-reactive IgG and Unequal Strain-specific Neutralizing Antibodies in Naïve and Previously Infected Recipients. 2021. doi: 10.1101/2021.06.19.449100.

10. Premkumar L, Segovia-Chumbez B, Jadi R, Martinez DR, Raut R, Markmann A, et al. The receptor binding domain of the viral spike protein is an immunodominant and highly specific target of antibodies in SARS-CoV-2 patients. Sci Immunol. 2020;5(48). Epub 2020/06/13. doi: 10.1126/sciimmunol.abc8413. PubMed PMID: 32527802; PubMed Central PMCID: PMCPMC7292505.

11. Lumley T. survey: analysis of complex survey samples. R package version 4.0 ed2020.

12. R Core Team. R: A language and environment for statistical computing. Vienna, Austria: R Foundation for Statistical Computing; 2019.

13. Yan D, Zhang X, Chen C, Jiang D, Liu X, Zhou Y, et al. Characteristics of Viral Shedding Time in SARS-CoV-2 Infections: A Systematic Review and Meta-Analysis. Frontiers in public health. 2021;9. doi: 10.3389/fpubh.2021.652842. PubMed PMID: 33816427.

14. Kissler SM, Fauver JR, Mack C, Olesen SW, Tai C, Shiue KY, et al. SARS-CoV-2 viral dynamics in acute infections. medRxiv. 2020. doi: 10.1101/2020.10.21.20217042.

15. Burgess S, Ponsford M, Gill D. Are we underestimating seroprevalence of SARS-CoV-2? BMJ (Clinical research ed). 2020;370. doi: 10.1136/bmj.m3364. PubMed PMID: 32883673.

16. Faustini S, Jossi S, Perez-Toledo M, Shields A, Allen J, Watanabe Y, et al. Detection of antibodies to the SARS-CoV-2 spike glycoprotein in both serum and saliva enhances detection of infection. medRxiv : the preprint server for health sciences. 2020. doi: 10.1101/2020.06.16.20133025. PubMed PMID: 32588002.

17. Lumley SF, Wei J, O’Donnell D, Stoesser NE, Matthews PC, Howarth A, et al. The duration, dynamics and determinants of SARS-CoV-2 antibody responses in individual healthcare workers. Clin Infect Dis. 2021. Epub 2021/01/06. doi: 10.1093/cid/ciab004. PubMed PMID: 33400782; PubMed Central PMCID: PMCPMC7929225.

18. Barzin A, Schmitz J, Rosin S, Sirpal R, Almond M, Robinette C, et al. SARS-CoV-2 Seroprevalence among a Southern U.S. Population Indicates Limited Asymptomatic Spread under Physical Distancing Measures. mBio. 2020;11(5). doi: 10.1128/mBio.02426-20. PubMed PMID: 32994333.

19. Bajema K, Wiegand R, Cuffe K, Patel S, Iachan R, Lim T, et al. Estimated SARS-CoV-2 Seroprevalence in the US as of September 2020. JAMA Intern Med. 2021;181(4). doi: 10.1001/jamainternmed.2020.7976. PubMed PMID: 33231628.

20. NC Department of Health and Human Services. COVID-19 Dashboard - Cases: NC Department of Health and Human Services; 2020 [April 29, 2021]. Available from: https://covid19.ncdhhs.gov/dashboard/cases.

